# Changes of the Commensal Microbiome during Treatment are Associated with Clinical Response in Nasopharyngeal Carcinoma Patients

**DOI:** 10.1101/2020.02.27.20028647

**Authors:** Tingting Huang, Justine Debelius, Alexander Ploner, Xiling Xiao, Tingting Zhang, Kai Hu, Zhe Zhang, Rensheng Wang, Weimin Ye

**Affiliations:** Department of Medical Epidemiology and Biostatistics, Karolinska Institutet, Stockholm, Sweden; Department of Radiation Oncology, The First Affiliated Hospital of Guangxi Medical University, Nanning, P. R. China; Radiation Oncology Clinical Medical Research Center of Guangxi, Nanning, Nanning, P. R. China; Department of Otolaryngology-Head & Neck Surgery, First Affiliated Hospital of Guangxi Medical University, Nanning, P. R. China

**Author notes:** Address correspondence to Weimin Ye and Rensheng Wang. Weimin Ye and Rensheng Wang contributed equally to this work. Author order was determined by consortium policy.

## Abstract

The human microbiome has been suggested to be involved in the regulation of response to anticancer therapies. However, little is known regarding changes of commensal microbes in cancer patients during radiotherapy and whether these changes are associated with response to treatment. We conducted a prospective, longitudinal cohort with sixty-two newly diagnosed nasopharyngeal carcinoma (NPC) patients who were scheduled for radiotherapy-based treatment. Nasopharyngeal swabs were collected longitudinally before radiotherapy, during radiotherapy, and after radiotherapy. The nasopharyngeal microbiome was assessed using 16S rRNA amplicon sequencing. All patients were followed up to 24 months to define an early or late clinical response. We demonstrated the beta-diversity of the nasopharyngeal microbiome showed temporal changes throughout treatment. The magnitude of changes was stably and significantly different between the early and late responders. The temporal microbial networks among NPC patients with early response differed significantly from those with late response. Seven amplicon sequence variants (ASVs) mapped to *Corynebacterium* were lost during treatment. Twenty-eight abundant ASVs differed by patients’ responses throughout treatment. Among them, 10 ASVs differed between the early responders and late responders before getting any treatment and the difference was consistent along the radiotherapy course. This study addressed the temporal changes of the nasopharyngeal microbiome in NPC patients during radiotherapy and suggested a significant association with clinical response. The subject-specific changes of the nasopharyngeal microbiome might serve as a potential predictor for clinical response to radiotherapy.

**Importance:** The human microbiome has been suggested to be involved in the regulation of response to anticancer therapies. However, little is known regarding changes of commensal microbes in cancer patients during radiotherapy and whether these changes have an impact on response to treatment. In this longitudinal study of nasopharyngeal carcinoma patients, we demonstrate that the temporal changes of the nasopharyngeal microbiome in NPC patients during radiotherapy-based treatment and suggest a significant association with patients’ clinical response. We identify 28 abundant amplicon sequence variants differed significantly between the early and late responders throughout treatment. Among them, 10 are consistently differed by patients’ responses. These subject-specific changes might serve as a potential predictor for clinical response to radiotherapy. To the best of our knowledge, this is the first example that the commensal microbiome may influence the response to radiotherapy-based treatment in cancer patients.

## Introduction

The human commensal microbiome has been suggested to be involved in both carcinogenesis and in the regulation of response to anticancer therapies (1-4). Accumulating evidence suggests that the microbiome can substantially affect the effectiveness of chemotherapy and immunotherapy such as gemcitabine and immune checkpoint inhibitors (5, 6). Accordingly, there is a growing interest in identifying and targeting these microbes in the anti-cancer treatment. However, little is known regarding whether or how the microbiome may regulate the response to radiotherapy, which is critical to the realization of its potential (2, 3, 7). In part, this is due to a lack of well designed longitudinal studies looking at the relationship between microbes and response to radiotherapy is seldom studied (2, 3).

To address this knowledge gap, we conducted a prospective study to characterize the longitudinal patterns of the commensal microbiome among cancer patients during radiotherapy-based anti-cancer treatment and investigated whether the temporal changes of the microbiome might be associated with patients’ clinical response. Nasopharyngeal carcinoma (NPC) is a malignant disease and endemic mainly in southern China and Southeast Asia (8). Radiotherapy is the essential mainstay of curative-intent treatment for the non-disseminated disease. Intensity-modulated radiation therapy (IMRT) contributes to the high 5-year survival rate of 68-80%, together with chemotherapy (9, 10). However, both local and distant failure after a first-line therapy remain key challenges which call for identification of novel indicators and modulators of clinical response. Hence, we prospectively collected nasopharyngeal swab samples during radiotherapy from a hospital-based NPC-patient cohort in southern China where NPC incidence rates exceed 10-15 per 100 000 person-years, about twenty times the rate in the western world (8). We hypothesized that the temporal change of nasopharyngeal microbiome during radiotherapy might be associated with NPC patients’ clinical responses.

## Results

A total of 445 nasopharyngeal microbial profiles, corresponding to 39 NPC patients, were analyzed. Thirty-six (92.3%) patients had advanced disease (stage III to IV_B_) at diagnosis. All of the NPC patients were treated with radiotherapy, 35 (89.7%) received concurrent chemotherapy including 25 (64.1%) who received induction chemotherapy or adjuvant chemotherapy. Twenty-seven patients (69.2%) with CR at the first clinical check-up were defined as early responders; the remaining 12 NPC patients achieved CR within 24 months of follow-up were defined as late responders (Supplementary Table S1).

### NPC patients’ clinical responses are associated with the temporal changes of between-sample diversity of nasopharyngeal microbiome during radiotherapy

The overall pattern of beta-diversity of the nasopharyngeal microbiome was visualized in PCoA projections for both unweighted and weighted UniFrac distance (Supplementary Figure S1). In both PCoA projections, samples of early responders were relatively dissimilar with the samples of late responders along PC2 (explained 5.88% of the variation in the unweighted projection; and 12.93% of the variation in the weighted projection), while similar along PC1 (13.23% in the unweighted, and 29.50% in the weighted) and PC3 (3.59% in the unweighted, and 7.84% in the weighted). By performing volatility analysis, the global variation (as an average) along PCs as a measure of longitudinal temporal volatility is presented in Figure 1. We saw a consistent shift along with the PC1 space with sampling time in both unweighted and weighted panels, regardless of clinical response. In PC2 panels, distinct separations between early and late responders were observed throughout treatment, whereas the temporal trajectories were relatively stable in each group. Moreover, in PC3 space, a moderate separation between the trajectories of response groups was found in the unweighted panel while the trajectories in the weighted panel showed fluctuated and gradual directional changes throughout treatment regardless of response status. The changed volatilities found in the last few sampling time-points have a larger standard deviation compared to the previous time-points, which might be partially explained by low sample numbers. Patient compliance with sample collection varied.

**Figure 1.**
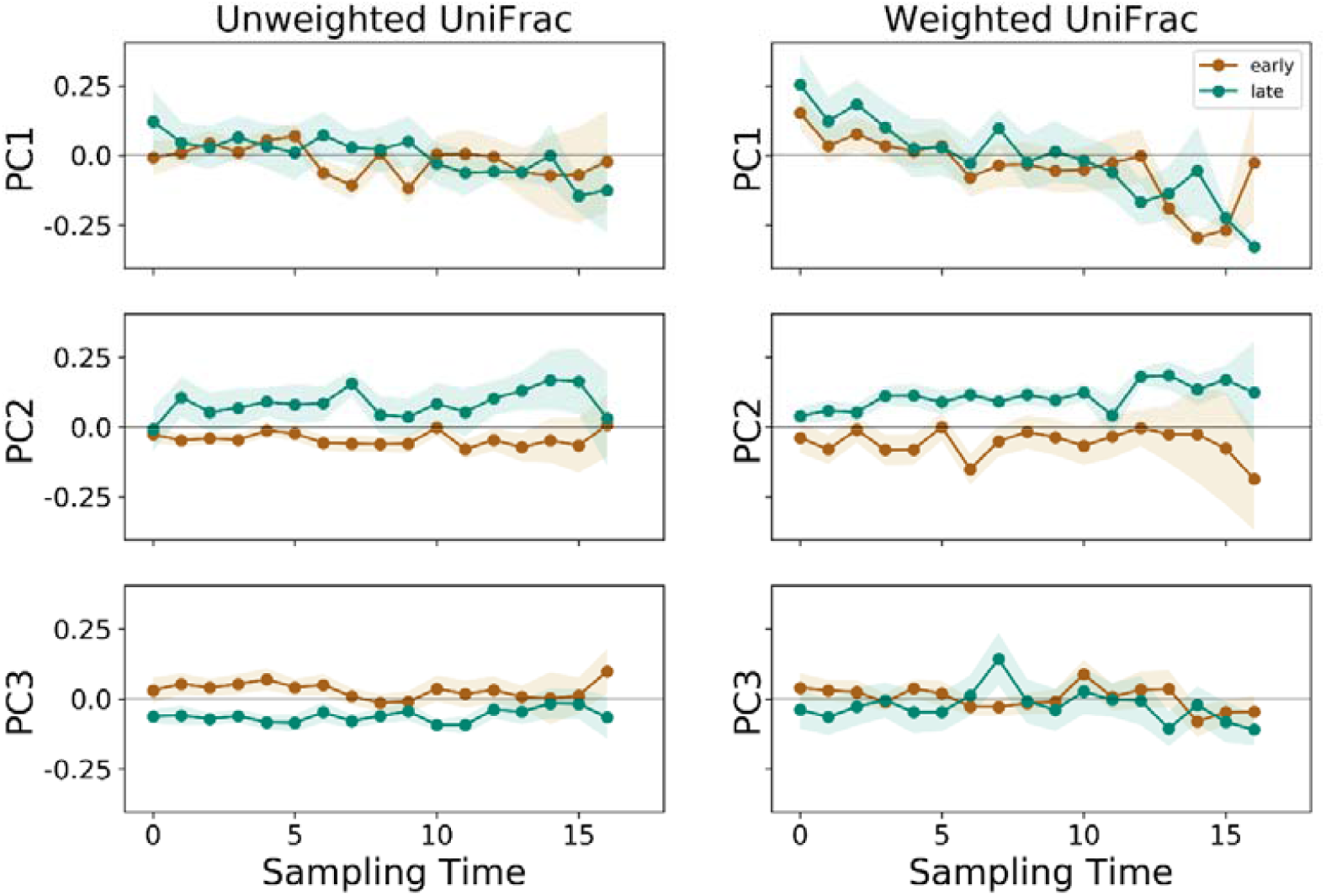
Normalized volatility of the nasopharyngeal microbiome in 39 patients during a 7-week treatment course. The longitudinal pattern of the nasopharyngeal microbiome of 39 patients over 7-week treatment was explored by Principle Coordinate Analysis (PCoA) projection along principle coordinate1 (PC1), PC2 and PC3 with regard to sampling time. The volatility analysis was used to visualize the mean distances based on UniFrac distance matrices in each PC space. The left of each panel reflects mean volatility of all individual trajectories referred to the unweighted UniFrac distance matrix, and the right side referred to the weighted UniFrac distance matrix. The consecutive numbers on the bottom indicate the sampling time points (0 = before radiotherapy). The trace represents the mean and the shaded is the standard deviation of each sampling time point. The curves in each PC space are marked by response status (a brown curve showing the early responders, a green curve showing the late responders).

We measured the rate of change in beta-diversity differences over radiotherapy in terms of the weighted UniFrac step length Δ-wUF and tested whether Δ-wUF changed over treatment and in response to the response status of NPC patients. We found statistically significant evidence for subject-specific differences (random effects) in baseline step length (intercept) as well as a change in step length over sampling time (slope, *P*-value = 0.0005) from linear mixed effect models (Supplementary Table S2). However, the across-subject average step length (fixed effect) appeared stable in Figure 2A, with no evidence for increasing or decreasing dynamic change (*P*-value = 0.4609). Adding the clinical response status improved the mixed effect model significantly (*P*-value = 0.0152). The best-fitting model including clinical response status retained stable for average step lengths for both early and late responders, with larger average step lengths for early responders compared to late responders (Figure 2B). Individual trajectories varied considerably between subjects, shown in Figure 2C.

**Figure 2.**
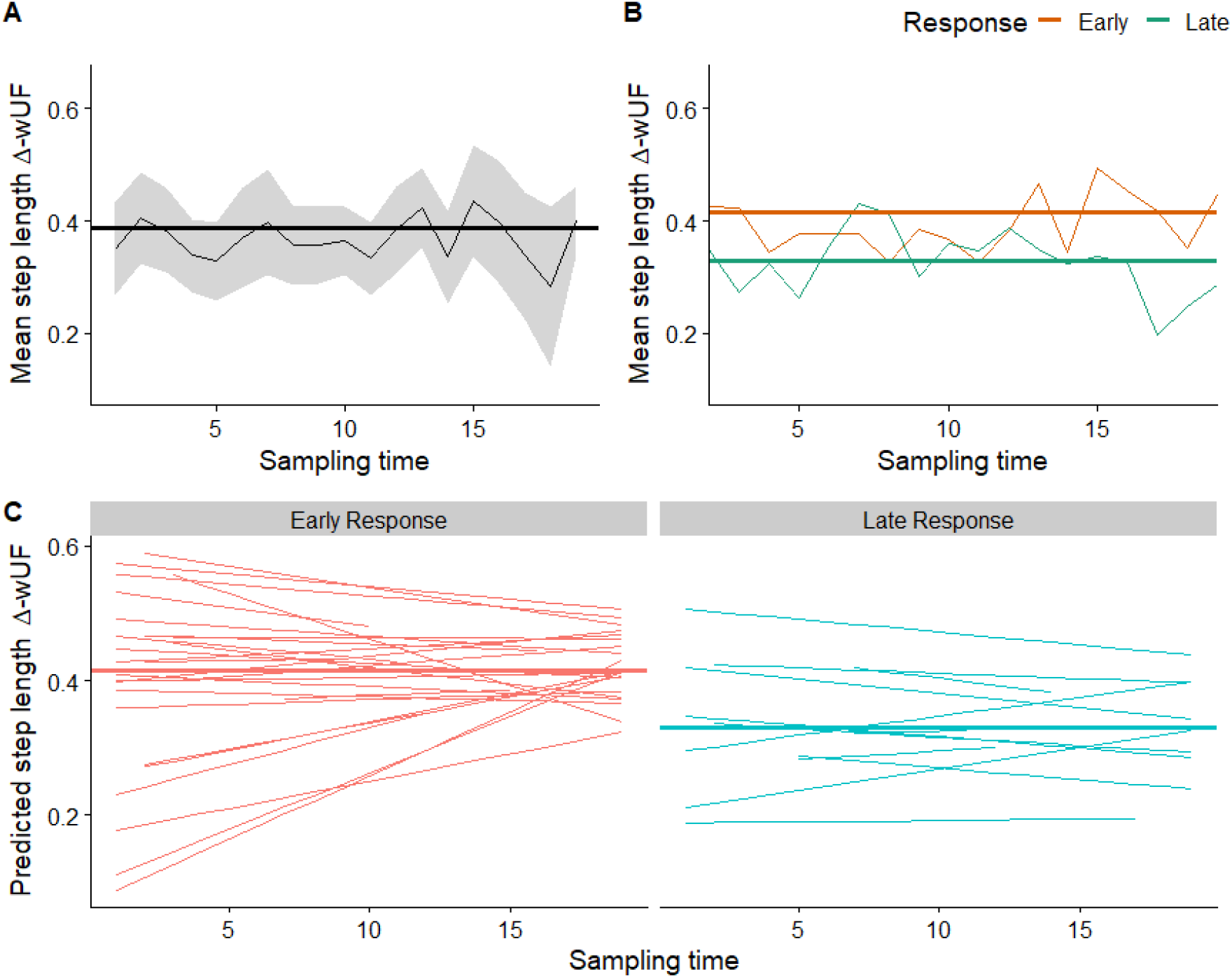
Group-wise and individual Δ-wUFs during treatment reflect the rate of change in the community. The weighted UniFrac step length Δ-wUF of an individual’s trajectory through nasopharyngeal microbial space was modeled as a function of treatment occasion (sampling time) via linear mixed effect models (LMEs), including within-individual random effects. The consecutive numbers on the bottom of each panel indicate the sampling time points. We explored (Panel A) the average step length across all individuals and (Panel B) average step length by clinical response. The moving average is shown as a thin line with shaded pointwise 95% confidence intervals; the thicker line is the estimated average step length (fixed effect) from the mixed model which takes into account the individual-level variations in step length over sampling time. We also explored the predicted individual step length trajectories for early and late responders (Panel C). The horizontal axis indicates the sampling time during treatment and the vertical axis the mean step length Δ-wUFs.

### NPC patients’ clinical responses are associated with the temporal trajectories of the abundant microbial features

Seventy-three ASVs were identified as the abundant subset for feature-based analyses. A Procrustean randomization test and Mantel test based on the Bray-Curtis distance indicated a well concordance between the abundant subset and the full dataset (PROTEST: correlation = 0.9669, *P*-value = 0.001, 999 permutations; Mantel test: correlation = 0.9506, *P*-value = 0.001, 999 permutations; Supplementary Figure S2). NMIT evaluates interdependence networks, which summarized the temporal trajectories of the abundant subset over treatment of each NPC patient into a single distance. The NMIT distances differed between clinical response groups. We observed that the interdependence networks among patients with early response statistically significantly differed from those with a late response, and no statistically significance differences found within response groups (PERMANOVA: *P*-value = 0.014; permdisp: *P*-value = 0.315; 999 permutations). In Figure 3, the networks of the late responders varied widely, forming a spread ellipsoid-like cluster, compared to the early responders who formed a smaller and more constrained ellipsoid-like cluster. These clusters are dissimilar along PC1 and PC3, but relatively similar along PC2.

**Figure 3.**
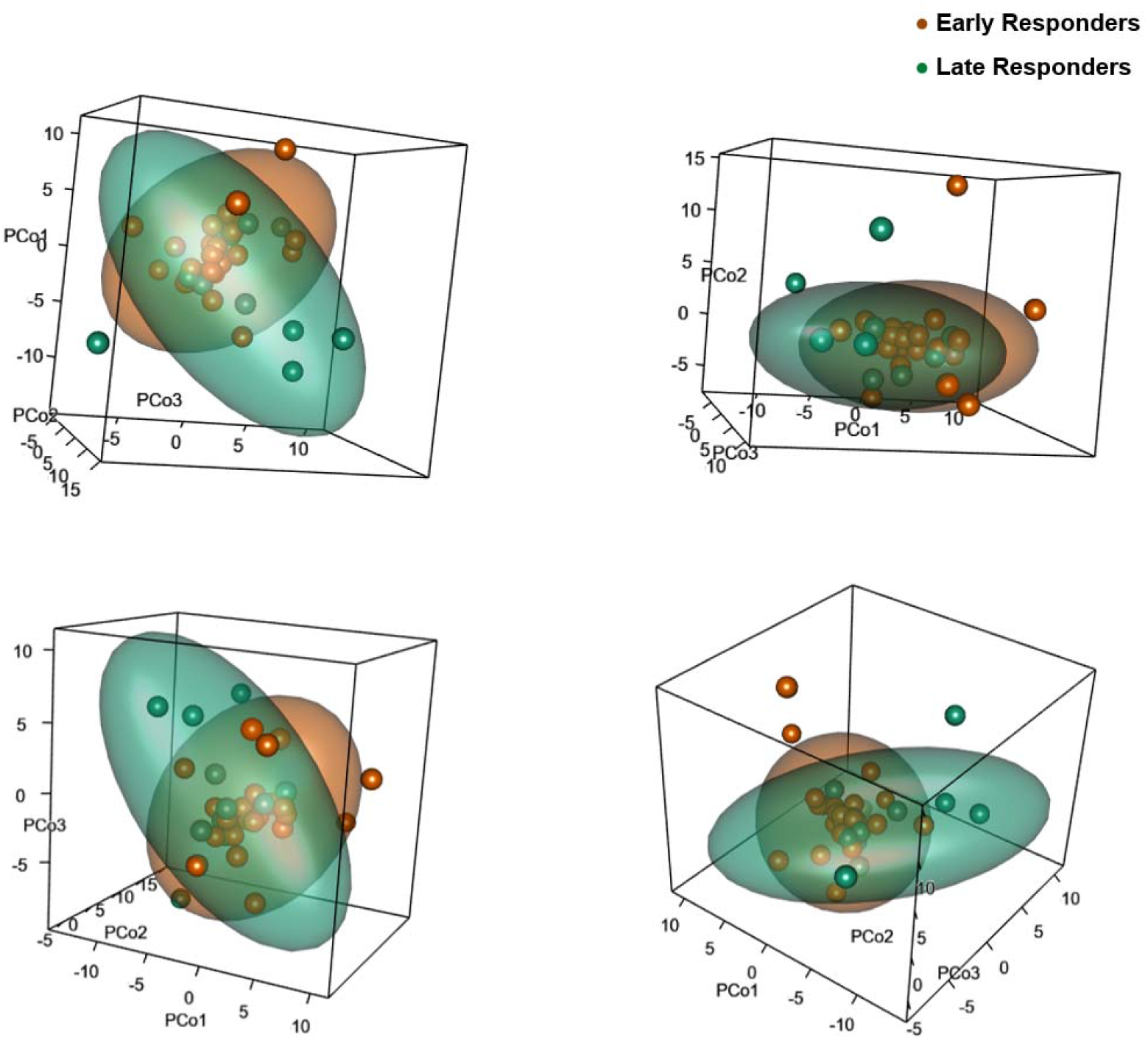
Differences in the interdependence networks of nasopharyngeal microbiome between the early and late responders. Non-parametric microbial interdependence test (NMIT) was used to determine longitudinal sample similarity as a function of temporal microbial composition networks over treatment. The similarities of nasopharyngeal microbial networks among NPC patients were visualized by Principle Coordinate Analysis (PCoA) projection. The first 3 axes in PCoA explain 15.03% of the variation in total (PCo1=5.67%, PCo2=4.91%, PCo3=4.45%). Each dot in the PCoA space is an indicator representing the temporal microbial network of one NPC patient. Brown dots are the early responders, green the late responders. The cloud representing the minimum-volume ellipsoid covers 80% of the data of each group. Panels A-D reflect different orientations of the same 3-dimention plot (screenshots from a 3-dimension PCoA projection).

We investigated longitudinal changes in the abundant ASVs across all samples using a modified ANCOM-test. We identified seven abundant ASVs where more than 80% of ASV ratios were temporally correlated (Supplementary Figure S3, panel A), all members of genus *Corynebacterium*. There was a decrease in the relative abundance of all seven ASVs showed during treatment while most other ASVs remained stable over treatment course (Supplementary Table S3; and Figure S3, panel B to E).

To quantify ASV-based longitudinal differences between early and late responders, we applied SS-ANOVA. We identified 28 out of 73 abundant ASVs as differing significantly between early and late responders over treatment course, listed in Table 1 (*P*-value < 0.05, 1500 permutations). Notable that, among these significant ASVs, 10 differed between the early responders and late responders before getting any treatment, and remained consistently different during radiotherapy. Additionally, there were 9 ASVs that were similar between response groups during the first one-third of treatment and became different until the completion. In contrast, 5 ASVs differed between groups at the beginning and was converged during treatment.

**Table 1.** Twenty-eight abundant ASVs were detected with significant differences between early and late responders during a 7-week treatment course.

| ASV-ID | Taxonomy<sup>a</sup> | Time intervals<sup>b</sup> |  | Permutative<sup>c</sup> |
| --- | --- | --- | --- | --- |
|  |  | Start | End | P-value |
| gBurkh.6364 | g__Burkholderia | 7 | 20 | 0.005 |
| gBurkh.39c8 | g__Burkholderia | 8 | 20 | 0.007 |
| gCandi.9a10 | g__Candidatus Rhodoluna | 14 | 20 | 0.009 |
| oBacil.0094 | o__Bacillales | 8 | 20 | 0.011 |
| gAcine.32b4 | g__Acinetobacter | 7 | 20 | 0.011 |
| gAcine.efe8 | g__Acinetobacter | 18 | 20 | 0.011 |
| gEnhyd.0ef3 | g__Enhydrobacter | 10 | 20 | 0.018 |
| gStrep.3ed3 | g__Streptococcus | 1 | 17 | 0.020 |
| gBrevu.1285 | g__Brevundimonas | 1 | 19 | 0.024 |
| gCoryn.ca4d | g__Corynebacterium | 8 | 20 | 0.029 |
| gBurkh.f802 | g__Burkholderia | 14 | 20 | 0.044 |
| gBurkh.ee2f | g__Burkholderia | 7 | 20 | 0.049 |
| fMethy.cbae | f__Methylobacteriaceae | 5 | 11 | 0.017 |
| gCoryn.1650 | g__Corynebacterium | 1 | 12 | 0.026 |
| fBrady.ebba | f__Bradyrhizobiaceae | 6 | 20 | 0.029 |
| gCoryn.5ba9 | g__Corynebacterium | 1 | 13 | 0.043 |
| gTherm.b26c | g__Thermus | 1 | 20 | 0.003 |
| fCaulo.a200 | f__Caulobacteraceae | 2 | 20 | 0.003 |
| gTherm.94db | g__Thermus | 1 | 19 | 0.003 |
| gTherm.087b | g__Thermus | 1 | 20 | 0.004 |
| gAcine.4f58 | g__Acinetobacter | 1 | 20 | 0.005 |
| gRalst.edc7 | g__Ralstonia | 1 | 20 | 0.006 |
| gAcine.4c95 | g__Acinetobacter | 1 | 20 | 0.009 |
| gAcine.dfae | g__Acinetobacter | 1 | 20 | 0.013 |
| gCoryn.ff32 | g__Corynebacterium | 1 | 12 | 0.016 |
| gStaph.ab0b | g__Staphylococcus | 1 | 14 | 0.021 |
| gCoryn.0838 | g__Corynebacterium | 1 | 15 | 0.025 |
| gRalst.a529 | g__Ralstonia | 4 | 20 | 0.041 |

<sup>a</sup>: Taxonomy assignment. “g\_\_” refers to genus; “f\_\_” refers to family; “o\_\_” refers to order.
<sup>b</sup>: Time intervals with significant difference; number refers to sampling time (1 = before radiotherapy, 20 = end of radiotherapy). <sup>c</sup>: 1500 permutations.

For better visualization of the temporal trajectories of the difference between the early and late responders, the log2-fold changes of the 73 abundant ASVs estimated by SS-ANOVA were plotted as a heatmap and clustered into three groups (Cluster A to C) (Figure 4, and Supplementary Table S4). We found 22 ASVs (30.1%) in Cluster A, which were more prevalent among the early responders while 12 of them were significantly different between response groups with an increased relative abundance along with treatment course. Among the significant ASVs, 4 were members of genus *Burkholderia* (gBurkh.39c8, gBurkh.6364, gBurkh.f802, and gBurkh.ee2f), 2 were genus *Acinetobacter* (gAcine.efe8, and gAcine.32b4), and the rest were members of genus *Corynebacterium* (gCoryn.ca4d), *Streptococcus* (gStrep.3ed3), *Brevundimonas* (gBrevu.1285), *Enhydrobacter* (gEnhyd.0ef3), *Candidatus Rhodoluna* (gCandi.9a10), and unspecified order Bacillales (oBacil.0094). In Cluster B (n=37), most of the ASVs had relatively small difference between response groups and were found no statistically significant difference across treatment. Four ASVs showing significant difference across treatment (assigned to genus *Corynebacterium* (gCoryn.1650, gCoryn.5ba9), unclassified family Methylobacteriaceae (fMethy.ebba), and unclassified family Bradyrhizobiaceae (fBrady.cbae)). For Cluster C, 14 ASVs were more among the late responders compared to the early responders with dynamic trajectories (increased, decreased, and relatively stable in differece) over treatment. Twelve of them found statistically significant which were the members of genus *Acinetobacter* (gAcine.4f58, gAcine.4c95, and gAcine.dfae), genus *Thermus* (gTherm.b26c, gTherm.94db, and gTherm.087b), genus *Ralstonia* (gRalst.edc7 and gRalst.a529), genus *Corynebacterium* (gCoryn.ff32, gCoryn.0838), genus *Staphylococcus* (gStaph.ab0b), and unspecified family Caulobacteraceae (fCaulo.a200).

**Figure 4.**
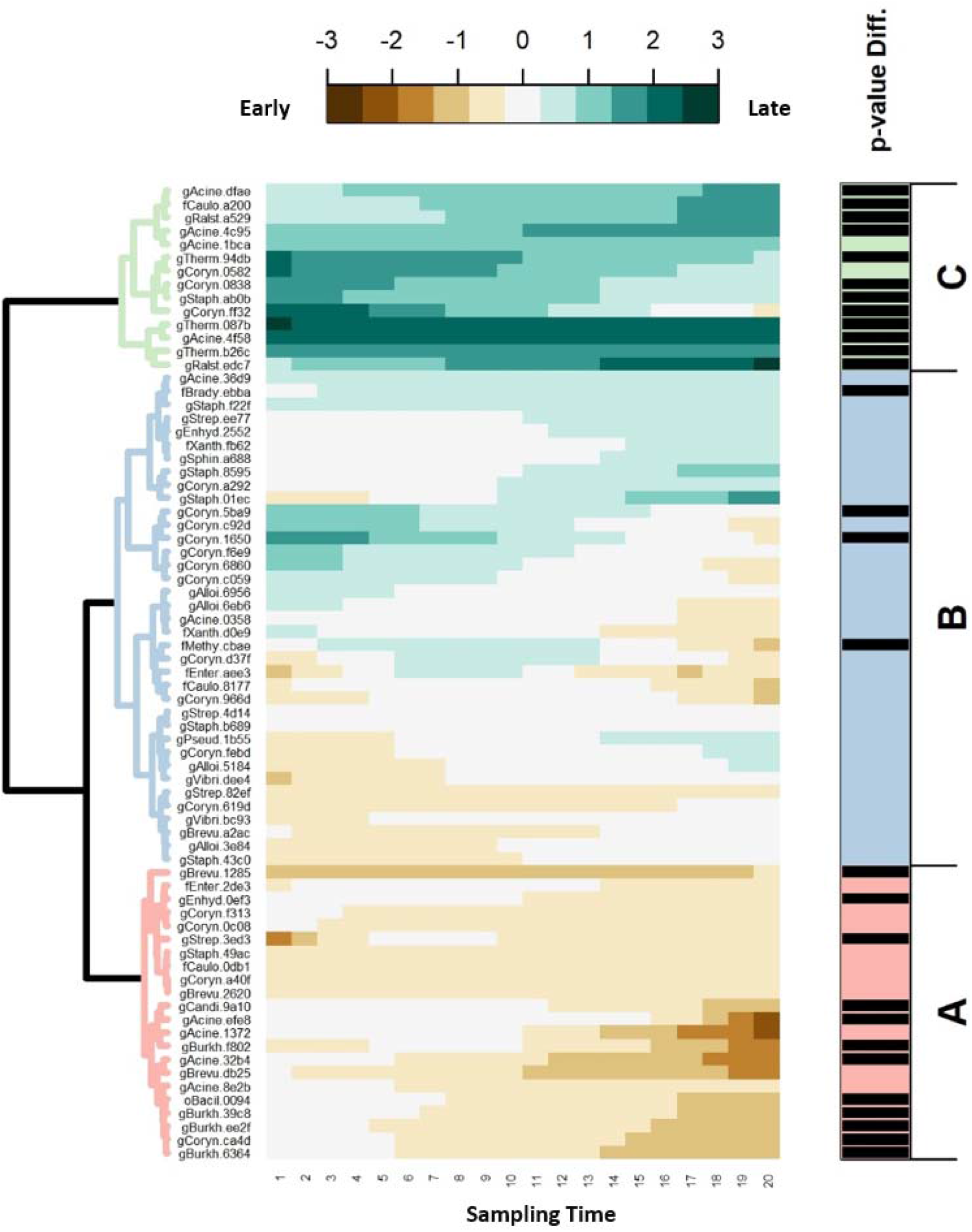
The predicted log2-fold change trajectories of 73 abundant ASVs between the early and late responders. The predicted log2-fold change trajectories of 73 amplicon sequence variants (ASVs) between the early responders and late responders generated from SS-ANOVA were grouped into hierarchical clusters using Ward’s D2 and displayed as a heatmap. Each raw in the heatmap represents the trajectory across treatment of each ASV. The consecutive numbers on the bottom of heatmap indicate the sampling time points (1 = before radiotherapy, 20 = end of radiotherapy). ASVs with higher abundance in early responders are brown; those with more abundance in late responders are teal. Three clusters are marked as A to C. The vertical bars on the right side indicate whether the trend in difference across treatment is statistically significant (*P*-value < 0.05 for the black bars, otherwise not).

## Discussion

In the present study, we have addressed temporal changes of the commensal microbiome in response to radiotherapy-based anticancer treatment among NPC patients for the first time. To be specific, the temporal changes in between-sample diversity of the nasopharyngeal microbiome and the abundant microbial features are associated with NPC patients’ clinical responses. The longitudinal study design provided a comprehensive view of microbial diversity, an effective way for assessing the microbial correlation networks and an opportunity to reveal the degree of change of the abundant features between time points during the observation period (11). Notably, the subject-specific changes showed in our study might suggest a possibility of using the nasopharyngeal microbiome as a predictor for clinical response.

We observed that the UniFrac distances used for measuring beta-diversity qualitatively (unweighted) and quantitatively (weighted), on average, showed temporal changes throughout treatment (volatilities along PC1 explaining the largest amount of within-subject changes). Previous studies also described the progressive alteration in the oral microbiome during radiotherapy among NPC patients (12), and the change in gut microbiome in patients after pelvic radiotherapy (13). Remarkably, we found distinct and consistent separations in volatility regarding patients’ clinical responses along the PC2, which suggests that a global difference between the early and late responders might have existed before treatment and stay relatively stable afterward until the completion of radiotherapy. We further demonstrated that the magnitude of longitudinal global changes of the nasopharyngeal microbiome was significantly different between response groups, namely the average step length of the early responders was statistically, significantly larger than that of the late responders. However, the magnitude of changes appeared constant within each group along with treatment. These findings challenge the Anna Karenina principle, which suggests the positive prognosis is associated with community stability (14). It may implicate bacterial resilience to radiotherapy as a feature of delayed response. It is reasonable to imagine that such a community might be protective in some way against both radiation-induced perturbations on the microbiome as well as the host.

In the feature-based analysis, we observed similar characteristics in the NMIT analysis: the temporal changes in microbial networks of NPC patients over treatment were statistically significantly different regarding their clinical responses. The difference observed between groups was not due to a large degree of dispersion within groups. By looking at individual abundant ASV, genera known to dominate the human upper aerodigestive tract: *Corynebacterium, Staphylococcus, Acinetobacter*, and *Streptococcus* (15, 16), were also found dominant in our study. We found a significant and consistent loss of *Corynebacterium* (including 7 distinct ASVs) over treatment. Oral *Corynebacterium* was reported previously to decrease the risk of head and neck cancer, although the study did not include NPC (17). Members of the genera are also normally, temporally stable in Chinese adults (18).

Additionally, we observed that 28 out of 73 abundance ASVs were significantly different in abundance between the early and late responders during treatment. Notably, 10 ASVs primarily found in Cluster C differed between groups before the initiation of radiotherapy-based treatment and consistently differed until the completion. These results correspond to the characteristics observed in the diversity-based analyses above. Moreover, genera *Ralstonia* and *Thermus* have been previously identified as reagent contaminants (19). We found consistent signals for 4 ASVs from these genera (Ther-b26c, Ther-087b, Rals-edc7, and Rals-a529) between response groups, despite sample randomization across plates. These organisms were at very low abundance or absent in early responders, but present in late responders. Thus, the finding is unlikely solely due to contamination. Members of these extremophile genera are known to be radiation- or ROS-resistant, which suggests a potentially plausible biological mechanism for their inclusion in these communities (20). These findings suggest a possibility that these ASVs might be putative indicators of patients’ response to radiotherapy-based treatment.. However, it is also important to note that some ASVs assigned to the same genus behaved differently, such as members of the genus *Acinetobacter* (gAcine.32b4 and gAcine.efe8 in Cluster A, compared to gAcine.4f58, gAcine.4c95, and gAcine.dfae in Cluster C), which may reflect niche specialization within the same genus (11).

While the results suggest a clear relationship between changes in the microbiome and treatment response, several open questions remain. First, previous studies in the literature focused mainly on potential associations between the commensal microbiome and the radiotherapy-induced side effects and comorbidities among cancer patients (12, 21-24), and rarely investigated the impact of microbiome on radiotherapy efficacy except a pilot study with a small sample size (n=3) which reported a relationship between the response to radiotherapy and the gut microbiome in pediatric cancer patients (25). It is, however, difficult to compare our findings directly with most of the published data. Replication studies will be needed to determine if the microbiome can be used as a reliable indicator of treatment response in NPC. In addition, we acknowledge that the majority of our study subjects were NPC patients with stage III to IVb diseases (more than 90%) who were administrated concurrent chemo-radiotherapy (platinum-based) as recommended by clinical guidelines, and the analyses were not adjusted by therapeutic strategies due to a concern of low statistical power. The response to platinum-based chemotherapy has been reported to be modulated by gut microbiota, which influences the inflammation, immunity, and metabolism systematically (2, 26). Our analyses could not distinguish the contributions from radiotherapy and chemotherapy, separately. However, we considered that the nasopharyngeal microbiome might be affected mainly and heavily by radiotherapy with high energy X-ray in the nasopharynx. Last but not the least, some researchers suggest that microbiome affects the response to radiotherapy by modulating the damage-associated molecular pattern signals, however, the essential knowledge is yet deficient and the underlying mechanism is yet to be explored (2, 4, 27). Therefore, additional work is warranted to replicate our findings among similar study populations; and to address potential mechanisms, such as immune regulation and potential interaction between the nasopharyngeal microbiome and systemic immunotherapy.

In conclusion, this study demonstrated the temporal changes of the nasopharyngeal microbiome in NPC patients during radiotherapy-based treatment and suggested a significant association with clinical response. By focusing on NPC, a type of cancer where radiotherapy is the most common treatment, we hypothesize that the relationship between the microbiome and clinical response may hold true for other diseases and anatomical locations. These results shed new light on a possibility that the commensal microbiome may influence the response to radiotherapy-based anticancer treatment in cancer patients, and call for larger longitudinal studies with long-term follow-up to understand the underlying mechanisms.

## Materials and Methods

### Study design and setting

This prospective study recruited newly diagnosed NPC patients in the First Affiliated Hospital of Guangxi Medical University, Guangxi Province located in southern China, between 2014 and 2015. The study was approved by the Ethical Review Committee of the First Affiliated Hospital of Guangxi Medical University, China, and the Regional Ethical Review Board in Stockholm, Sweden. Written informed consent was obtained from all patients. A total of 76 NPC patients were recruited and 62 were enrolled in this study. Details are given in Supplementary Data (Supplementary Method S1 and Figure S4).

All patients were treated according to the standards of clinical practice (details are given in Supplementary Method S2). All patients received the first clinical check-up at 3 months after the completion of radiotherapy and were followed up for 24 months. Patients who achieved a complete response (CR) in the first check-up were classified as early responders and patients who achieved CR between the first and the end of follow-up were designated late responders.

### Sample collection and processing

In total 870 nasopharyngeal swabs were collected from 62 NPC patients. A protocol including enzymatic lysis and bead beating for DNA extraction was used. The indexed library targeting the V3-V4 hypervariable regions of the 16S rRNA gene was prepared using samples with sufficiently high-quality DNA. Extraction controls, PCR amplification controls, and bacterial reference controls were included in each step. After quality and quantity checking, a total of 526 16S rRNA-based libraries and 85 control libraries were sequenced by Beijing Genomics Institute (BGI, Wuhan, China), using a 300-bp paired-end strategy on Illumina Miseq according to the manufacturer’s instructions. Details are given in Supplementary Data (Supplementary Method S3 and S4, and Figure S4).

### Data denoising and pre-processing

Raw sequences were quality filtered and denoised using *deblur* to generated amplicon sequence variants (ASVs) in QIIME 2 (v. 2019-10) (28-31). A phylogenetic tree was built using fragment insertion into the August 2013 Greengenes 99% tree and taxonomic assignments using a naïve Bayesian classifier trained against the same reference (32-34). The dataset was filtered to remove samples without valid clinical information (4 individuals with 56 samples), ASVs assigned to mitochondrial rRNA (35), and samples with fewer than 1,500 reads (25 samples). Finally, 445 samples derived from 39 NPC patients were retained, with around 3 million high quality reads assigned to 9,320 ASVs (33 phyla, 107 classes, 192 orders, 336 families, and 724 genera). Between-sample diversity (beta-diversity) was estimated after samples were rarefied to 1,500 reads per sample (36). Feature-based analyses were performed using a representative subset of ASVs with at least 0.1% relative abundance in at least 10% of samples, as abundant ASVs (n=73). Two samples were removed due to the absence of abundant ASVs, retaining 443 samples. The concordance between the main ASVs dataset and the abundant ASVs subset was tested. Details are given in Supplementary Data (Supplementary Method S5 and Figure S5).

### Statistical analysis

In this study, the analyses of global patterns were mainly focused on beta-diversity using unweighted and weighted UniFrac distance (37, 38). Principal Coordinate Analysis (PCoA) projections were used to visualize between-sample difference (39). The longitudinal pattern of nasopharyngeal microbiome along each principle coordinate (PC) was visualized using volatility analysis in QIIME 2 2019.10 (*q2-longitudinal*) (40). The weighted UniFrac distance matric was further used to measure the rate of change which differed over treatment. The change in an individual community between two sequential treatment occasions was quantified as the weighted UniFrac distance between successive samples (Δ-wUF = wUF_(t)_ – wUF_(t-1)_), corresponding to the weighted Unifrac step length of an individual’s trajectory through nasopharyngeal microbial space. Δ-wUF was modeled as a function of treatment occasion (sampling time) via linear mixed effect models (LMEs, considering treatment, temporal and inter-individual effects) with the *lme4* package (v. 1.1-19) in R 3.5.1 (41). Differences between nested models were tested via likelihood ratio tests, while Akaike’s information criterion (AIC) was used to compare general models.

In the analyses of feature-based patterns, Non-parametric Microbial Interdependence Test (NMIT) was used to determine longitudinal sample similarity as a function of temporal microbial composition network over treatment (42). We further eliminated three patients who had less than five successive samples, retaining 434 samples of 36 patients, to make the analysis robust and efficient. In brief, the Spearman correlations between any pair of 73 abundant ASVs were computed based on their relative abundance; distances between any two of patients’ correlation matrices were calculated based on a Frobenius norm; PCoA was used to visualize the similarity among patients. The difference of microbial networks between response groups was tested by the permutation-based extension of multivariate analysis of variance (PERMANOVA) test (43) and the multivariate dispersion test (permdisp) (44). NMIT, PERMANOVA, and permdisp were carried out by QIIME 2 2019.10, and PCoA was plotted in R 3.5.1. On the other hand, the longitudinal patterns of the 73 abundant ASVs individually were also analyzed. The temporal relationship for the abundant ASVs was evaluated using a modified analysis of composition of microbiomes (ANCOM)-based approach using Spearman correlation (45). A normalized ANCOM W statistic was calculated as the fraction of tests where the raw p-values were less than 0.05, and a normalized W of more than 80% was considered statistically significant. This test was applied to both the raw data, and the ratios between time points. Additionally, the longitudinal differences in abundance (log2-transformed) of abundant ASVs between early and late responders were modeled via Smoothing-spline ANOVA (SS-ANOVA) (46) using *metagenomeSeq* package (v. 1.24.0); the predicted changing trajectories of abundant ASVs were grouped into hierarchical clustering using Ward’s method (Ward’s D2) and displayed as a heatmap using *Heatplus* package (v. 2.32.1) (47) in R.3.6.2.

### Data availability

Raw sequencing data and metadata are deposited in ENA under accession XXXXX.

## Data Availability

Raw sequencing data and metadata are deposited in ENA under accession XXXXX. More details can be achieved by contacting the corresponding authors.

## Acknowledgements

We acknowledge funding from the Swedish Cancer Society (2016/510 to W. Ye) and the Swedish Research Council (2015-02625, 2015-06268, 2017-05814 to W. Ye); the National Natural Science Foundation of China (81360405 to R. Wang) and the Guangxi (China) Science and Technology Program Project (GK AD17129013 to R. Wang); the Natural Science Foundation of Guangxi Medical University for Junior Scientists (GXMUYSF201203 to T. Huang). T. Huang was also partly supported by a grant from the China Scholarship Council (201408450018).

We would like to thank the patients recruited in this study, and the clinical staff in the Department of Radiation Oncology in the First Affiliated Hospital of Guangxi Medical University (Nanning, P. R. China). We are grateful to Dr. Zhe Zhang, Dr. Xiaoying Zhou, Dr. Xue Xiao, and their research group for lab support.

T. Huang had full access to all of the data in the study and takes responsibility for the integrity of the data and the accuracy of the data analysis. T. Huang, Z. Zhang, R. Wang, and W. Ye conceived and designed the study. T. Huang, R. Wang, and W. Ye received funding. T. Huang, T. Zhang, and K. Hu recruited patients, collected samples and follow-up information. T. Huang and X. Xiao performed laboratory assays. Z. Zhang, R. Wang, and W. Ye contributed resources. T. Huang, J. Debelius, and A. Ploner analyzed the data, prepared figures. T. Huang, J. Debelius, A. Ploner, R. Wang, and W. Ye interpreted the results. T. Huang, J. Debelius, and A. Ploner wrote the manuscript. All authors edited the manuscript and approval of the final draft.

On behalf of all authors, the corresponding authors declared that they have no potential financial and non-financial competing interests.

